# Baseline profile of intrinsic cytokines predicting prognosis of chronic hepatitis B patients responding to HBV therapeutic vaccinations

**DOI:** 10.1101/2022.04.18.22273944

**Authors:** Xiang Jin, Hongyu Jia, Gan Zhao, Fan Yu, Huan Cai, Lishan Yang, Sheng Jiang, Feifei Yang, Jie Yu, Shuang Geng, Weidong Zhao, Guodong Yu, Xiaoli Zhang, Jueqing Gu, Chanyuan Ye, Shanyan Zhang, Yingfeng Lu, Heng Liu, Huangli Meng, Jimin Zhang, Yida Yang, Bin Wang

## Abstract

**Objective:** To explore relevant biomarkers in chronic HBV (CHB) infected individuals, and whether their presence can be related to the prognosis of CHB (i.e., used as a prediction tool) and used as inclusion and exclusion criteria in clinical trials.

**Methods:** Thirty-four (34) cytokines and chemokines were analyzed in the baseline plasma of 130 chronic HBV infected patients and were matched with the clinical outcomes of these patients regarding to their responses to anti-HBV treatment by a mathematic model based on the Boolean method. A retrospective analysis was implemented to establish the prediction model, and a perspective analysis was performed to verify the prediction efficacy.

**Results:** Through analyzing 34 cytokines and chemokines in the baseline plasma of 130 chronic HBV infected patients by Boolean methods, we generated a predicting model successfully capable of screening out therapy non-responded patients. In this prediction model, six cytokines, including IL-8, IL-10, IL-17, IL-1RA, IFN-α, IL-18, defined as expressed or not-expressed, contributed to 21 possibilities, every of which predicts a clinical outcome. The model was verified in a separate chronic HBV infected population database, which included 76 patients, with 100% responders and 50% who are not responded to the immunotherapy identified.

**Conclusions:** The prediction model can be used to screen CHB patients as the inclusion incorporated into HBV clinical design and practice. By screening out inappropriate participants in clinical trials, therapy response rate may rise and lead to a more homogeneous responding population. For Boolean method which requires continuous iteration, more accurate prediction models will be established with more homogeneous data. This is very helpful for revealing the reason why certain CHB individuals can be functionally cured and others were not. The method may also have great potential and possible applications for other immunotherapies in the future.

**Significance of this study:** *What is already known about the subject?:*

a. Chronic hepatitis B virus (CHB) infection can be controlled while rarely cured, or functionally cured. The exact reason why certain CHB individuals can be functionally cured and others were not, regarding to different treatment strategies, remains unclear.
b. Lack of relevant immunological biomarkers are often to blame clinical failures in immunotherapeutic treatments, particularly for the hepatitis B virus (HBV) therapeutic vaccination, since such trials use virological parameters as inclusion and exclusion criteria of patients, but seldom more relevant immunological biomarkers.

*What are the new findings?:*

a. Using patterns of cytokines, instead of single cytokines, to present CHB individuals’ immune status can help discovering the prognosis of their responses or not response to HBV therapeutic vaccination.
b. By utilizing the model, we predicted 10 patients out of 10 who were sensitive to the anti-HBV immunotherapy and 33 out of 66 who were not, in a distinct CHB population, and verified the predicting efficacy.

*How might it impact on clinical practice in the foreseeable future?:*

a. Immune status, presented by different patterns of cytokines/chemokines, might be used as an in/exclusion criteria in clinical trials to select a more appropriate treatment for CHB individuals.
b. By screening out inappropriate participants in clinical trials, therapy response rate may rise and lead to a more homogeneous responding population. For Boolean method which requires continuous iteration, more accurate prediction models will be established with such more homogeneous data. This is very helpful for revealing the reason why certain CHB individuals can be responsive to the treatments and toward the functionally cured and others could not.

## INTRODUCTION

Chronic Hepatitis B virus (HBV) infection continues to be a significant burden in public health, with 257-270 million people seropositive of hepatitis B surface antigen worldwide in 2015, and the number is still increasing[1]. Over 86 million people, more than 6.1% of the population in China, were estimated to live with Chronic HBV in 2018, according to the Polaris Observatory Collaborators[2, 3]. Meanwhile, the upgrade and the availability of antiviral drugs and therapeutic vaccines against the virus are developing[4]. Unlike antiviral drugs, which effectively inhibit HBV replication though may lead to resistant viral variants in the long-term treatment, therapeutic vaccines are designed as alternative approaches alone or as add-on treatments to increase therapeutic efficacy and avoid antiviral drug resistance[5].

Some therapeutic vaccines have been proved to inhibit the disease deterioration and its potential of virus seroconversion[6]. Though the determinants of seroconversion of HBV infection remain not fully understood, the immune response is known to be involved in treatment outcomes [7]. However, lack of relevant immunological biomarkers are often to blame clinical failures in immunotherapeutic treatments, particularly for the HBV therapeutic vaccination, since such trials use virological parameters as inclusion and exclusion criteria of patients, but seldom more relevant immunological biomarkers. Cytokines are initially involved in host immune responses, while limited information would be acquired from a sole cytokine since they interact and regulate with each other directly or indirectly. Multiple cytokines assays of host immune response have been conducted to show a landscape of cytokine profile in a patient, and its efficacy in stratifying patients to therapy has been proved in different contexts [8, 9]. Finding a single biomarker directly linked to the outcome of immunotherapy was approved to be difficult due to the complex and intermingled connection of cytokine networks in different patients at various stages of disease progressions and the contribution of human leukocyte antigen (HLA) haplotypes.

Herein, using a Boolean method, we generated a prediction model which can rule out the nonresponse chronic hepatitis B (CHB) individuals to the HBV therapies by specific patterns of cytokine status, showing that the 6-cytokines, consisting of IL-8, IL-10, IL-17, IL-1RA, IFN-α, IL-18, can be used to stratify a subset of patients who were not sensitive to the immunotherapy before the therapy starts. These findings could help screen CHB patients for further treatment and promote a deeper understanding of HBV seroconversion determinants. In addition, the Boolean method applied here has been proved effective and worth promoting into more disease conditions or therapeutic approaches.

## RESULTS

Plasma samples from a total of 130 patients were collected for 34 immune biomarkers as the source data for establishing a prediction model. All cytokines/chemokines were detected to better characterize the immune status of each patient at an early stage and can be categorized into four groups as described in Materials and Methods. After all plasma concentrations of immune biomarkers were detected, we converted the status of each immune biomarker into binary classification, defined as Expressed (1) and Not-expressed (0) according to the cut-off, the detecting limit of the Luminex system, lower than what was defined as 0, otherwise defined as 1. In this way, the immune status of each patient can be summarized into a 34-digit serial number, as “0000…0000000”, “100100…0010001”, etc., as Figure 1A. Every 34-digit number represents a different panel of 34 combined cytokines encoded by Boolean operations on the network nodes. Each panel, characterized 34-digit number, was then defined as a criterion to distinguish the serological converted and unconverted patients. At weeks 0 (baseline), 22 cytokines were detected expressed or not expressed in all patients and were then deleted from the cytokine pools. At week 12, 18 cytokines were removed for the same reason.

**Fig 1.**
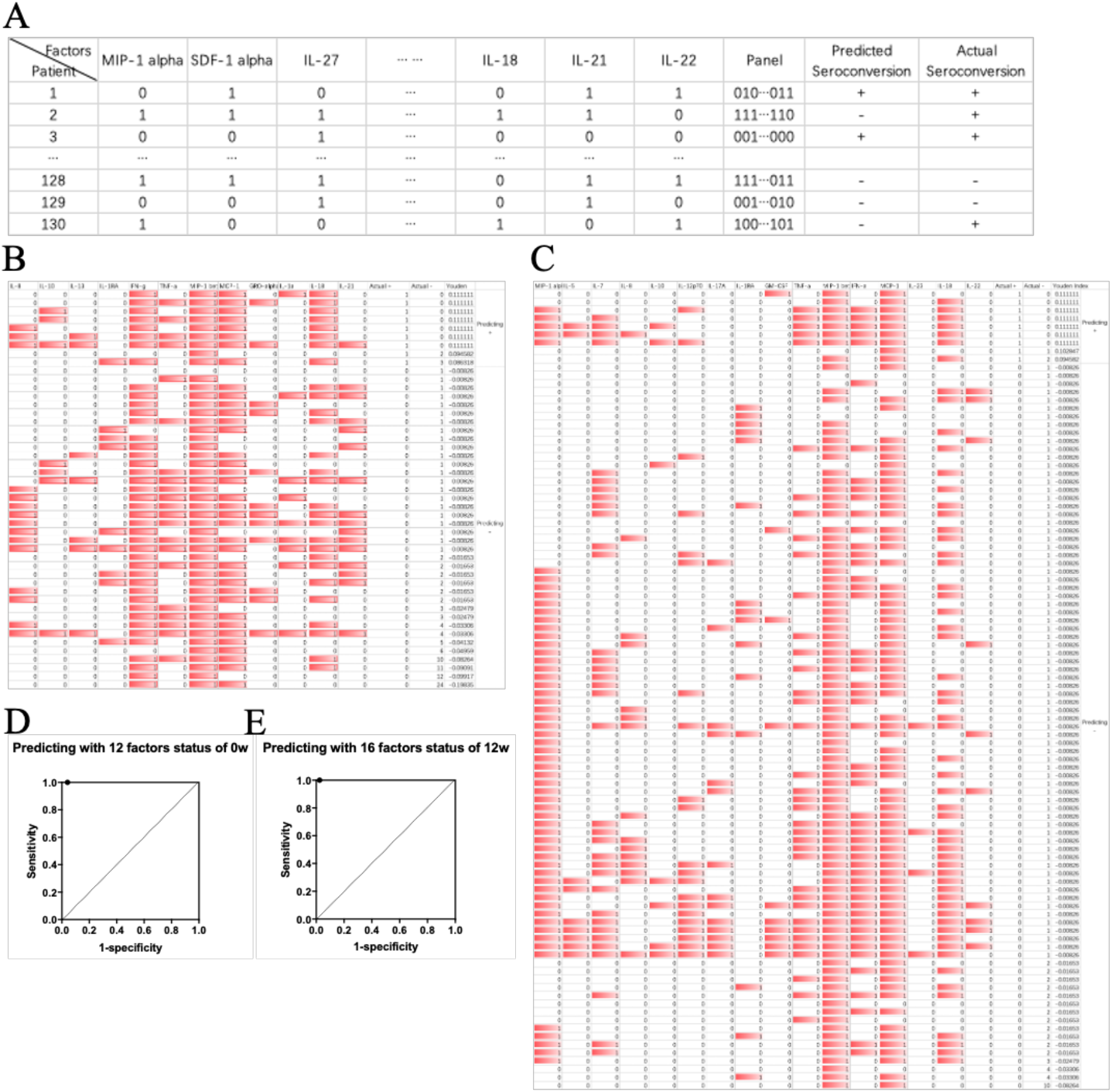
Multiple cytokine status analysis. **A:** Each cytokine was represented by the digit number “1” or “0”,meaning “on”(Expressed) or “off”(Non-Expressed) respectively according to its expression in patient’s plasma compared with cut-off. Each X-digits number(X=the number of factors involved) represents a panel in prediction matched to the actual situation. With a certain number of factors (immune biomarkers), the prediction model composed of the panels; **B:** The outcome of predicting the sero-conversion and non-conversion using 34 cytokines status in patient’s week 0 plasma, 12 of 34 cytokines remained; **C:** The outcome of predicting the sero-conversion and non-conversion using 34 cytokines status in patient’s week 12 plasma, 16 of 34 cytokines remained; **D** and **E:** Predicting ROC with 12 factors at week 0 and with 16 factors at week 12 respectively.

Consequently, the length of panel digit numbers was reduced from 34 to 12 and 16 at week 0 and week 12, respectively (Figure 1B, C). The 12- and 16-digit numbers at weeks 0 and 12, which included 47 and 98 immune states, respectively, herein defined as panels, were used for prediction model optimization in this study. Each panel was then evaluated by Youden Index (Figure 1B, C). Under this scheme, the Youden Index of these 12-cytokine panels is 0.9586, with sensitivity =1 and specificity =0.958678 (Figure 1D), which means all (100%) serological converted patients and 95.86% of non-converted patients were predicted before starting therapy according to their cytokine status at week 0. The Youden Index of this 16-cytokine panel was 0.9752, with sensitivity =1 and specificity =0.9752 (Figure 1E).

To achieve equivalent prediction efficacy while excluding less relevant immune biomarkers, we evaluated every single factor in prediction and parallelly removed each factor at a time from the whole panel to observe the influence of the removal. Not surprisingly, no single biomarker stood out as the driving force, while combining these biomarkers significantly improved prediction performance (see Figure 1B, C). In this step, we involved an overall model combined with both prediction models based on weeks 0 and 12. The Youden index of the overall model remained at 0.9338 (Figure 2E). With this overall model, multiple All-Minus-One cytokine prediction models were then iterated to determine the most relevant cytokine in predicting serological conversion, as shown in Figure 2F-H. Ultimately, a 6-cytokine model was discovered, including IL-8, IL-10, IL-17, IL-1RA, IFN-α, IL-18, the Youden Index of 0.7649, with sensitivity =0.8889 and specificity=0.876 (Figure 2I). This model comprises 21 panels through different combinations of the six cytokines’ expression statuses (Figure 2J).

**Fig 2.**
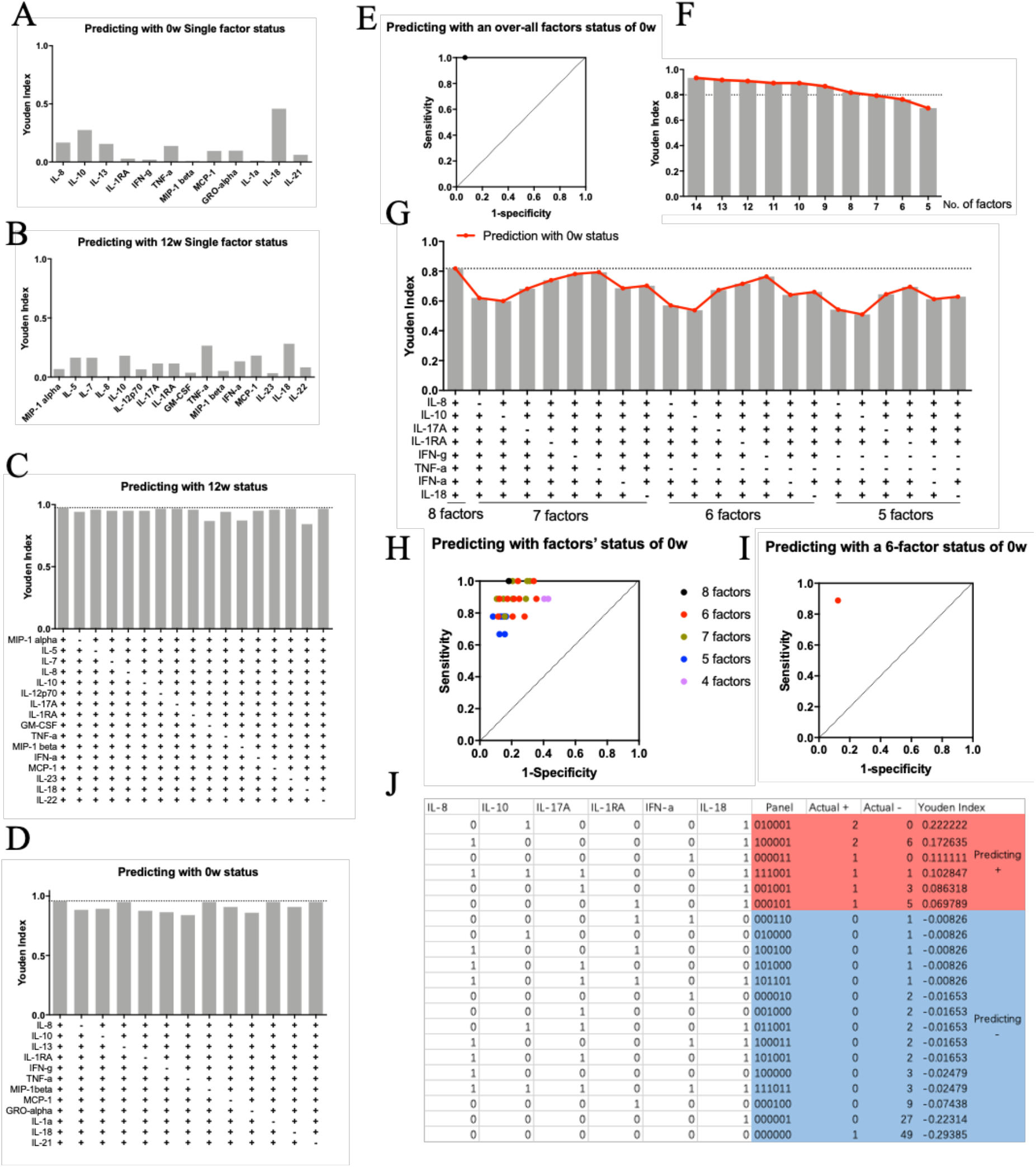
Generation of prediction model. **A-B:** Youden Index of Predicting sero-conversion and non-conversion with single cytokine status at week 0 and week 12,12 and 16 cytokines were included initially; **C-D:** Youden Index change of predicting sero-conversion and non-conversion when remove one cytokine off the initial included cytokines at week 0 and week 12; **E:** Evaluation of combined factors from week 0 and week 12, including MIP-lα, IL-7, IL-8, IL-10, IL-17A, IL-IRA, IFN-γ, TNF-α, MIP-β, IFN-α, MCP-1, IL-18, IL-21,IL-22, others were excluded for lower contribution to the prediction; **F:** Youden Index of All-minus-one cytokines; **G-H:** Youden index and ROC of multiple factors in prediction respectively; **I:** ROC of final 6-cytokine model; **J:** All panels in 6-cytokine model and its predicting outcomes.

After establishing the 6-cytokine model, we examined the levels of HBsAg of infected individuals, as the predicting outcome measurement, during the 72 weeks of therapy to validate the accuracy. Across the entire source data, it showed that the 6-cytokine model could predict 106 serological non-converted patients out of 121 actual non-converted patients and eight converted patients out of 9 actual converted patients based on baseline status. No significant difference was observed in patients who are not serological converted (Figure 3A-B), demonstrating that the predicting results were consistent with actual clinical outcomes and had great potential in screening out therapy insensitive patients.

**Fig 3.**
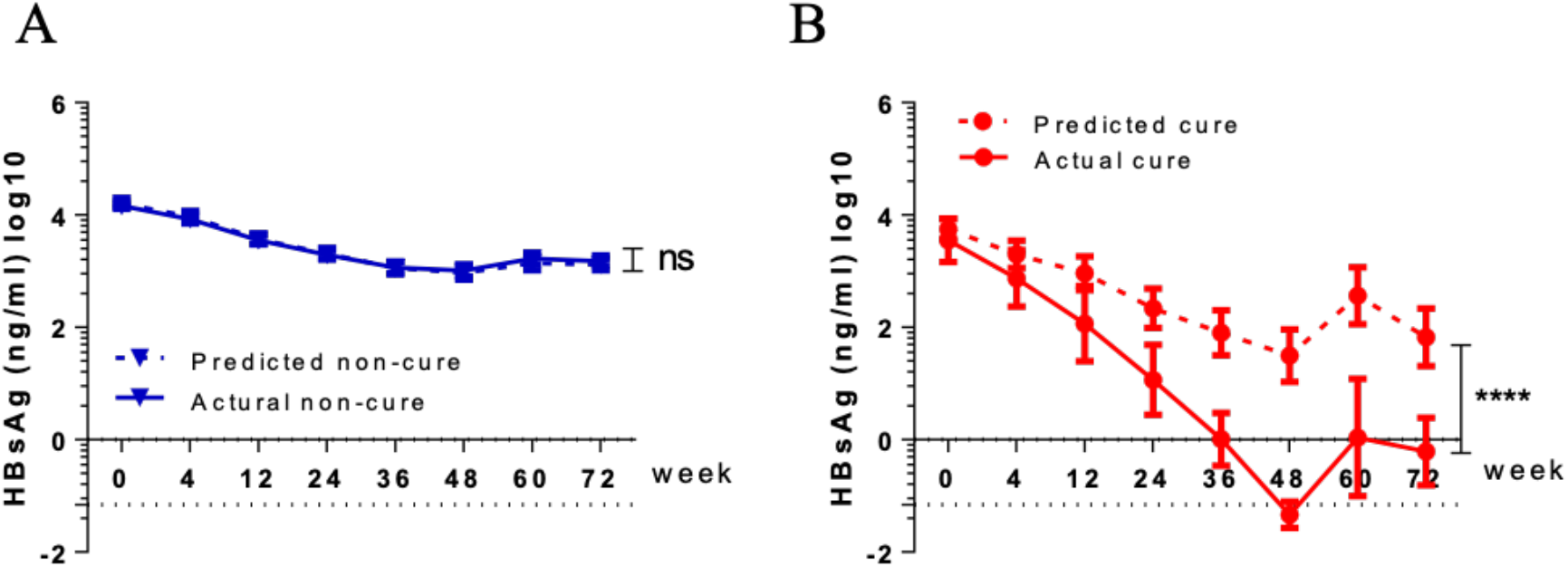
Predicting efficacy in virological data. The dynamic of HBsAg between predicting and actual status was compared according to the clinical outcomes during the treatment and follow-up. Non-conversion **(A)**, Sero-conversion **(B)**.

Finally, to illustrate the utility of our prediction model, we utilized this panel in predicting a separate 76-patient pool, which was not presented in our training data. All of the 76 patients were HBV chronically infected and had been on therapy. The immune biomarker status of all 76 patients at week 0 was detected and defined as described above. At week 0 (baseline), we correctly identified all 10 patients who were serologically converted during the therapy and 33 patients out of 66 not serological converted, when matching their clinical outcomes at week 72, consistent with the conclusion. The virological data were validated by matching the prediction outcomes afterward, shown in Figure 4.

**Fig 4.**
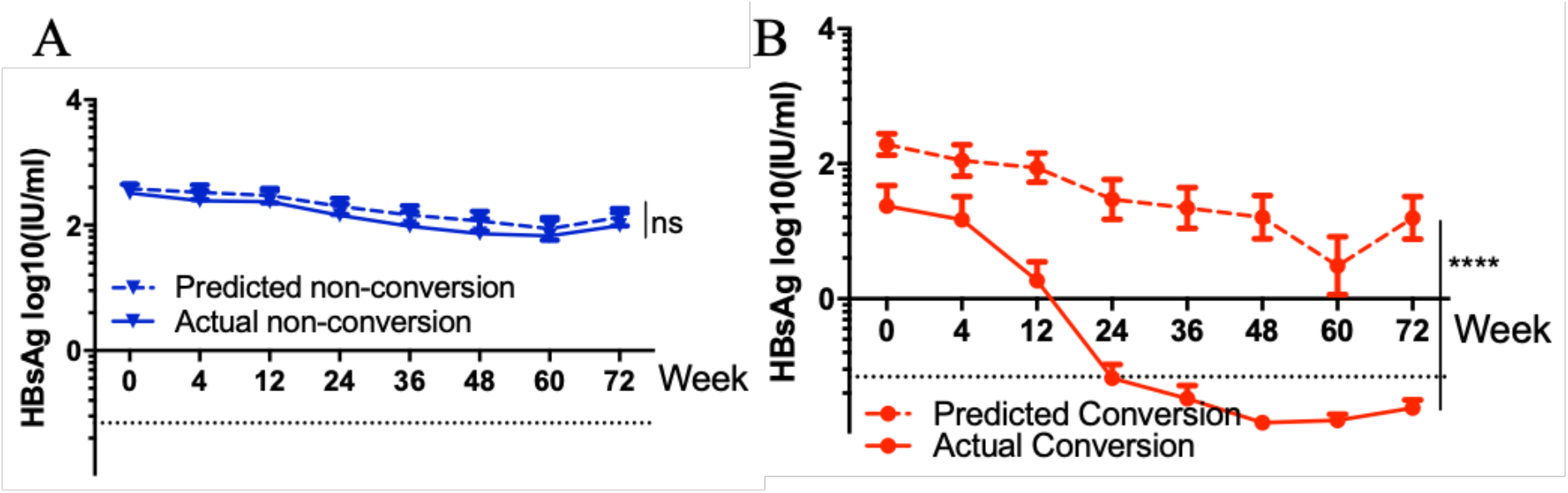
Verification of the predicting model efficacy. The 6-cytokine prediction model was utilized in another independent HBV chronic infected population, and the dynamic of HBsAg between predicting and actual status was monitored and then compared according to the clinical outcomes during the treatment and follow-up. Non-conversion **(A)**, Sero-conversion **(B)**.

## DISCUSSION

The selection of appropriate patients prior to HBV therapeutic clinical trials, including HBV therapeutic vaccine clinical trial, is essential for the clinical outcomes and financial expenditures and for patients to get beneficial treatments. The therapeutic vaccines reconstruct the immune response by overcoming immune tolerance. During the process, including the reactivation of the immune system to the virus, it has also been proved that the host immune status plays a crucial role[10]. Thus, analyzing the contribution of host factors is required to discover more of the determinants of immunotherapeutic efficacy. Cytokines and chemokines are involved in all immune reactions with interferences and feedbacks, implying that a single factor reveals much less information than their combination due to their complex interaction networks, similarly to our results (Figure 1B, C). In our study, to better characterize the host immune status, we selected 34 related cytokines and chemokines and then combined multiple cytokines/chemokines status as Expressed (1) and Not-expressed (0) to a serial number, representing a panel. Subsequently, we utilized a Boolean method by matching the host immune status (the panel) at baseline with the clinical outcomes during the 72 weeks after treatment and generated a prediction model for screening appropriate patients in further clinical studies. The results have shown that these six cytokines, IL-8, IL-10, IL-17, IL-1RA, IFN-α, IL-18, of all 34 cytokines, were detected and involved in differentiating patients who were unlikely to respond to the immunotherapy from patients were. By utilizing this model, we predicted 10 patients out of 10 who are sensitive to the therapy and 33 out of 66 who are not in another HBV chronic infected population, verifying the predicting efficacy.

The 6-cytokine panel includes IL-8, IL-10, IL-17, IL-1RA, IFN-α, IL-18, none of which alone is significantly related to the loss of HBsAg, while showing excellent capability predicting the outcomes in combination. In other studies, it has been found that all these cytokines are associated with HBV infection. IL-8, with its mitogenic and angiogenic functions, has been associated with tumors and chronic inflammatory diseases[11]. IL-10, secreted mainly from T cells with inhibition effects of various cytokines from T cells and monocyte/ macrophages, has been correlated with virus clearance during HBV infection[11]. IL-18 was initially discovered as an interferon-g-inducing factor, now is known as a potent pro-inflammatory cytokine involved in host defense and an immune activator[12]. In our study, patients infected with HBV with higher IL-18 levels are more likely to have HBsAg seroconversion and have also been shown relevant to seroconversion in other studies[13]. When predicting alone, IL-17, IL-1RA, and IFN-α showed limited efficacy in the model, while in other studies, clinical evidence has shown the association of these cytokines with HBV progression[11, 14, 15]. The reason why our study did not show these single cytokine/chemokine correlations to the outcomes may be the limitation of the sample size and our HBV infected patients unstratified with certain serum status. In addition, our results indicate that the combination of these cytokines, representing the immune status, explains much more than each of them.

Including in clinical trials, selecting appropriate patients is essential not only for the clinical outcomes and financial expenditures but for patients to get more appropriate treatments. Multiple predicting models have been generated and been successfully applied in complex biological networks[16-19], including Boolean methods[20]. During our process of generating this Boolean model, the immune status was generated, representing each patient to match the clinical outcomes, calculating the probability. In this way, based on a 130 patients pool, we figured out a 6-cytokine prediction panel suitable for the 130-patient pool itself, as well as another new 76-patient pool. The limitation here is, by using this model, we differentiated a subset of the patients who are unlikely to respond to the therapy while are unable to screen out patients who have better responses ideally. This may be associated with patients’ relatively low response ratio to the treatment. Larger sample size and more extensive cytokine/chemokine can be further iterated in this model for optimization.

In summary, based on serum cytokine/chemokine data of 130 HBV infected patients, we generated a 6-cytokine model in predicting patients who are not sensitive to the therapy at baseline, which can help rule out this subset of patients and improve the efficacy of the therapy. Further research on the model generation and cytokine status with a more significant number of patients are required for better performances, and this Boolean model generated by cytokine status might be promoted in other disease or therapeutic approaches.

## Material and Methods

### Study Samples and Characteristics of the Study Population

Plasma samples were used for cytokine/chemokine assays. Plasma was aliquoted, stored at −80 °C, and assayed for HBsAg. Samples used for the primary analysis had been taken on the day before treatment week 0.

Of all 130 patients included in the source database, all were aged between 18-65 and had never been treated with the following inclusion criteria: HBsAg (+), titer <10,000 IU/ml, HBeAg (+), ALT >2 x normal level, liver inflammation score > G2, and HBV DNA > 10^5^ copies/ml; 9 patients showed HBsAg seroconversion, and 121 patients did not, undergo a 72-week observation during anti-HBV therapies and follow-ups.

In addition, data of 70 CHB patients among 130 patients were from a study investigating the efficacy of immune tolerance-breaking therapy, treatments including adefovir dipivoxil (ADV) (Group 1), ADV+Interferon-α (IFN-α) (Group 2), and ADV+IFN-α+THRee Injections of Low-dose GM-CSF followed by one dose of the HBV Vaccine (THRIL-GM-vac) (Group 3). Seventy (70) patients completed the assigned treatment and follow-up period: 25 patients in ADV (Group 1), 22 patients in ADV+IFN-α (Group 2), 23 patients in ADV+IFN-α+THRIL-GM-vac (Group 3), with 0.0%, 4.5%, and 21.7% of patients of each group were serological converted, respectively (Figure 5). Detailed study results can be found in Manuscript Seroclearance of HBsAg in Chronic Hepatitis B Patients after a Tolerance-Breaking Therapy: GM-CSF, followed by Human HBV Vaccine.

**Fig 5.**
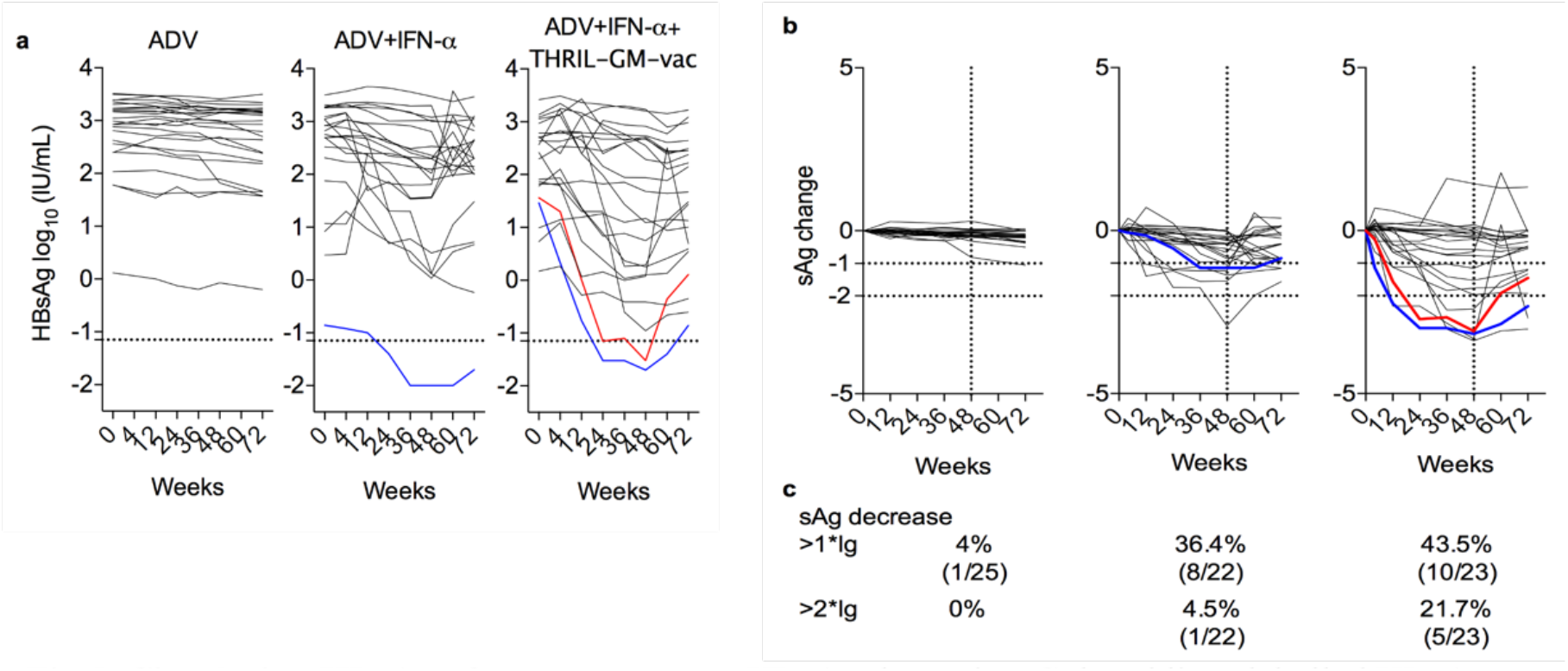
Circulating HBsAg after treatment. **a.** HBsAg dynamics. Colored lines labelled HBsAg seroclearance patients (HBsAg<0.07IU/mL, dash line), **b-c.** Individual normalized HBsAg dynamics by subtraction of baseline level. Horizontal dash lines represent 1-log and 2-log sAg decrease. Vertical dash line represents end of treatment at week 48.

### Luminex Cytokine/Chemokine multiplex assays

Thirty-four cytokines/chemokines were measured using a human MILLIPLEX MAP kit (Millipore), including four groups: chemokines (Eotaxin, GRO-α, IP-10, MCP-1, MIP-1α, MIP-1β, RANTES, SDF1-α), innate immune related cytokines (GM-CSF, IFN-α, IL-1β, IL-1α, IL-1RA, TNF-α, TNF-β), adapted immune related cytokines (IFN-γ, IL-2, IL-4, IL-5,IL-6, IL-7, IL-8, IL-9, IL-10, IL-12 p70, IL-13, IL-15, IL-17A, IL-18) and HBV related cytokines (IL-21, IL-22, IL-23, IL-27, IL-31).

Plates were read with a Luminex instrument (Luminex 200, Austin Luminex, USA) according to the manufacturer’s instructions. According to the manufacturer’s standard curves, each target’s concentration was analyzed using MILLIPLEX Analyst 5.1 software (Merck Millipore Darmstadt, Germany). The concentrations of various cytokines/chemokines were displayed as heat maps by Muti-Experiment Viewer 4 software (TM4 Group; Dana Farber Cancer Institute, Boston, MA).

### Boolean modeling

A Boolean method was utilized to generate a model to discover correlations between cytokines and final seroconversion. In brief, the status of each immune biomarker was defined as Expressed (1) and Not-expressed (0) according to the cut-off, the detecting limit of the Luminex system, lower than which was defined as 0, otherwise defined as 1. After the status transformation, a serial number consisting of 1 and 0s, also indicated as a prediction panel, was generated representing the immune status of each patient. One hundred and thirty serial numbers of all patients, including the HBsAg-seroconverted and non-converted, made up a Boolean states pool. Finally, by matching each serial number to the outcome of the patients, we calculated the possibility of each match to evaluate the validity of the prediction. To optimize the prediction model, the cytokines were stepwise removed to better performance. The Youden Index was used to define the effectiveness of the prediction model and calculated by sensitivity and specificity.

### Statistical analysis

The statistical analyses performed unpaired t-tests and two-way (ANOVA) analysis with Prism 7.0 software. Statistical significance was defined by P < 0.05.

### Ethics Statement

The clinical samples for this research were approved by the Research Ethics Committee of Huashan hospital, Fudan University. All participants where clinical samples originated from were enrolled in Huashan Hospital, Fudan University, and the First Affiliated Hospital, College of Medicine, Zhejiang University, all of whom signed informed consents. Relevant clinical trial had been registered on Chinese Clinical Trial Registry (https://www.chictr.org.cn), registered number as ChiCTR-TRC-13003254.

### Reporting guidelines

TRIPOD reporting guidelines[21]were used for this research.

## Supporting information

Supplemental Reporting Guideline Checklist

## Data Availability

All data produced in the present study are available upon reasonable request to the authors.

## Acknowledgements

This work was partly supported by the National Major Science and Technology Research Projects for the Control and Prevention of Major Infectious Diseases in China (2017ZX10202202, 2017ZX10202203007, 2013ZX10002001) to Drs. JM Zhang, HY Jia, LD Yang, and B.Wang, and the National Natural Science Foundation of China (81871640, 82172255) and Shanghai Municipal Science and Technology Major Project to B. Wang.

## Author Contributions

The prediction model was designed by BW, XJ, SG, and FY, and was generated by XJ. HYJ, JMZ, LDY managed the operation of relevant clinical trials. HC, LSY, FFY, JY, GDY, XLZ, JQG, CYY, SYZ, YFL, HL, HLM managed patients samplings, recordings and clinical analysis. XJ, FY, SJ, GZ, WDZ performed cytokines/chemokines experiments and analyzed data. XJ and BW wrote and edited the manuscript.

