## Supplemental Reporting Guideline Checklist for "Baseline profile of intrinsic cytokines predicting prognosis of chronic hepatitis B patients responding to HBV therapeutic vaccinations"

### Reporting checklist for prediction model development/validation.

Based on the TRIPOD guidelines.

| | | Reporting Item | Page Number |
|---|---|---|---|
| **Title** | | | |
| | #1 | Identify the study as developing and / or validating a multivariable prediction model, the target population, and the outcome to be predicted. | 1 |
| **Abstract** | | | |
| | #2 | Provide a summary of objectives, study design, setting, participants, sample size, predictors, outcome, statistical analysis, results, and conclusions. | 3 |
| **Introduction** | | | |
| | #3a | Explain the medical context (including whether diagnostic or prognostic) and rationale for developing or validating the multivariable prediction model, including references to existing models. | 4 |
| | #3b | Specify the objectives, including whether the study describes the development or validation of the model or both. | 5 |
| **Methods** | | | |
| Source of data | #4a | Describe the study design or source of data (e.g., randomized trial, cohort, or registry data), separately for the development and validation data sets, if applicable. | 13 |
| Source of data | #4b | Specify the key study dates, including start of accrual; end of accrual; and, if applicable, end of follow-up. | n/a |
| Participants | #5a | Specify key elements of the study setting (e.g., primary care, secondary care, general population) including number and location of centres. | n/a |

| | | | |
|---|---|---|---|
| Participants | #5b | Describe eligibility criteria for participants. | 14 |
| Participants | #5c | Give details of treatments received, if relevant | n/a |
| Outcome | #6a | Clearly define the outcome that is predicted by the prediction model, including how and when assessed. | 10 |
| Outcome | #6b | Report any actions to blind assessment of the outcome to be predicted. | 10 |
| Predictors | #7a | Clearly define all predictors used in developing or validating the multivariable prediction model, including how and when they were measured | 15 |
| Predictors | #7b | Report any actions to blind assessment of predictors for the outcome and other predictors. | n/a |
| Sample size | #8 | Explain how the study size was arrived at. | n/a |
| Missing data | #9 | Describe how missing data were handled (e.g., complete-case analysis, single imputation, multiple imputation) with details of any imputation method. | n/a |
| Statistical analysis methods | #10a | If you are developing a prediction model describe how predictors were handled in the analyses. | 16 |
| Statistical analysis methods | #10b | If you are developing a prediction model, specify type of model, all model-building procedures (including any predictor selection), and method for internal validation. | 15 |
| Statistical analysis methods | #10c | If you are validating a prediction model, describe how the predictions were calculated. | 5 |
| Statistical analysis methods | #10d | Specify all measures used to assess model performance and, if relevant, to compare multiple models. | 10 |
| Statistical analysis methods | #10e | If you are validating a prediction model, describe any model updating (e.g., recalibration) arising from the validation, if done | n/a |
| Risk groups | #11 | Provide details on how risk groups were created, if done. | n/a |
| Development vs. validation | #12 | For validation, identify any differences from the development data in setting, eligibility criteria, outcome, and predictors. | 10 |

**Results**

| | | | |
|---|---|---|---|
| Participants | #13a | Describe the flow of participants through the study, including the number of participants with and without the outcome and, if applicable, a summary of the follow-up time. A diagram may be helpful. | 14 |
| Participants | #13b | Describe the characteristics of the participants (basic demographics, clinical features, available predictors), including the number of participants with missing data for predictors and outcome. | 14 |
| Participants | #13c | For validation, show a comparison with the development data of the distribution of important variables (demographics, predictors and outcome). | 14 |
| Model development | #14a | If developing a model, specify the number of participants and outcome events in each analysis. | 6 |
| Model development | #14b | If developing a model, report the unadjusted association, if calculated between each candidate predictor and outcome. | n/a |
| Model specification | #15a | If developing a model, present the full prediction model to allow predictions for individuals (i.e., all regression coefficients, and model intercept or baseline survival at a given time point). | 10 |
| Model specification | #15b | If developing a prediction model, explain how to the use it. | 8 |
| Model performance | #16 | Report performance measures (with CIs) for the prediction model. | n/a |
| Model-updating | #17 | If validating a model, report the results from any model updating, if done (i.e., model specification, model performance). | 10 |

**Discussion**

| | | | |
|---|---|---|---|
| Limitations | #18 | Discuss any limitations of the study (such as nonrepresentative sample, few events per predictor, missing data). | 13 |

| | | | |
|---|---|---|---|
| Interpretation | #19a | For validation, discuss the results with reference to performance in the development data, and any other validation data | 13 |
| Interpretation | #19b | Give an overall interpretation of the results, considering objectives, limitations, results from similar studies, and other relevant evidence. | 13 |
| Implications | #20 | Discuss the potential clinical use of the model and implications for future research | 13 |
| **Other information** | | | |
| Supplementary information | #21 | Provide information about the availability of supplementary resources, such as study protocol, Web calculator, and data sets. | n/a |
| Funding | #22 | Give the source of funding and the role of the funders for the present study. | 16 |

The TRIPOD checklist is distributed under the terms of the Creative Commons Attribution License CC-BY. This checklist was completed on 15. April 2022 using https://www.goodreports.org/, a tool made by the EQUATOR Network in collaboration with Penelope.ai
